# MATERNAL DETERMINANTS OF ADVERSE NEONATAL OUTCOMES IN A RURAL DISTRICT HOSPITAL IN EAST AFRICA

**DOI:** 10.1101/2023.06.20.23291654

**Authors:** Adenike Oluwakemi Ogah, James Aaron Ogbole

**Author notes:** **Corresponding author:** Dr Adenike Oluwakemi Ogah, PhD, PhD, Department of Pediatrics and Child Health, School of Medicine, University of Zambia 1.

## Abstract

**Background:** Maternal predisposing factors to adverse birth outcomes are often times assumed to be similar in rural and urban settings. This assumption have led to many failed or failing interventions. This study investigated the maternal risk factors of adverse birth outcomes in a remote community and compared with existing literature of similar studies done in urban areas or developed settings.

**Subject and methods:** This was the baseline data of a prospective cohort study, carried out in Gitwe village, Rwanda, 2019. Healthy, 529 mother-singleton infant pairs were recruited consecutively from Gitwe district hospital.

**Results:** The burden of adverse neonatal outcomes of significance in this rural study (cesarean section delivery, low birth weight, small for gestational age and prematurity) were 38.8%, 10.6%, 21.4% and 4.9%, respectively. Significant (p<0.05) maternal characteristics associated with cesarean section delivery were obesity, high number of antenatal visits (>6), non-christian religion, university education, entrepreneurs, positive HIV status and short stature. Unmarried mothers were likely to produce LBW and preterm babies, while primips were prone to deliver SGA babies. The magnitude of adverse birth outcomes in this rural study was unexpectedly higher than what exist at and their drivers were not exactly the same as in urban settings and at national level.

**Conclusion and Recommendations:** The burden of adverse birth outcomes in this study was higher than that of several countries in the world. Therefore, mothers (not neglecting their marital, HIV, parity and religion status), residing in these rural areas should be priortised for health care interventions, in order to lower the short-and long-term effects of these adverse birth outcomes.

## Background

Adverse birth outcomes are measures of health at birth and their magnitude is dramatically decreasing in the past 40 years. However, there is still a large gap between developing and developed countries.^1^ Infants with one or more adverse birth outcomes are at greater risk for mortality and a variety of health and developmental problems. Adverse birth outcomes includes preterm birth, low birth weight, stillbirth, macrosomia, congenital anomaly, and infant/neonatal death.^1^ These adverse birth outcomes often require cesarean section delivery and contribute to more than 75% of neonatal deaths that occurred in the first weeks of life. Preterm birth is a live birth before 37 completed weeks of gestation and is the leading cause of neonatal mortality.^2^ Low birthweight is an infant, who weighed less than 2500 g at birth, regardless of gestational age.^3^

The overall prevalence of adverse birth outcomes is 25.7% in rural India,^4^ 15.61% in Zimbabwe,^5^ and 10.8% in rural Uganda.^6^ Historically, rural areas have had more healthcare problems than urban areas, most likely due to lower healthcare provision and utilization.^7^

Maternal factors associated with neonatal mortality, which is the extreme of adverse birth outcomes in the same study site (Gitwe District Hospital, Southern Province of Rwanda), as observed by Ndayisenga (2014) included grandmultigravida, positive HIV status, infrequent antenatal clinic visit and home delivery.^8^ Kibe et al. (2022) noted that cesarean section deliveries were disproportionately high among women of high socioeconomic groups, residence in Kigali city and attended at least four antenatal care visits. But cesarean section was significantly lower among multiparous mothers.^9^ In an hospital-based unmatched prospective case-control study, in Western Ethiopia, a predominantly rural region, the odds of developing adverse birth outcomes were 3.92 times higher in mothers, who had one-time ANC attendance than mothers who attended four and above times (2020).^10^ Vahid et al. (2023) observed in a retrospective study of 8888 pregnant mothers in Iran, those living in rural areas had a higher risk of developing anemia, preterm birth, post-term pregnancies, LBW, need for neonatal resuscitation, and NICU admission, but a lower risk of cesarean section, compared to those living in urban areas.^11^

Similarly, cesarean section rate was higher in urban (10.4%) than rural areas (3.8%) in Ahinkorah et al. (2022) multi-country study in sub-Saharan Africa. Wealth index (39.2%), antenatal care attendance (13.4%), parity (12.8%), mother’s educational level (3.5%), and health insurance subscription (3.1%) explained approximately 72% of the rural–urban disparities in caesarean deliveries. After conducting secondary analysis of the demographic and health surveys of 28 nations, these authors postulated that, if the child and maternal characteristics were levelled, more than half of the rural–urban inequality in adverse birth outcomes would be reduced^12^. For effective attenuation of adverse birth outcomes, data pertaining to determinants of adverse birth outcomes are important. The findings of this study will assist the health care policy makers and providers, in planning tailored interventions to improve the wellbeing of children and women in the rural areas.

## Concept of the study

Figure 1, shows the maternal determinants of adverse birth outcomes, that were investigated in this study.

**Fig. 1:**
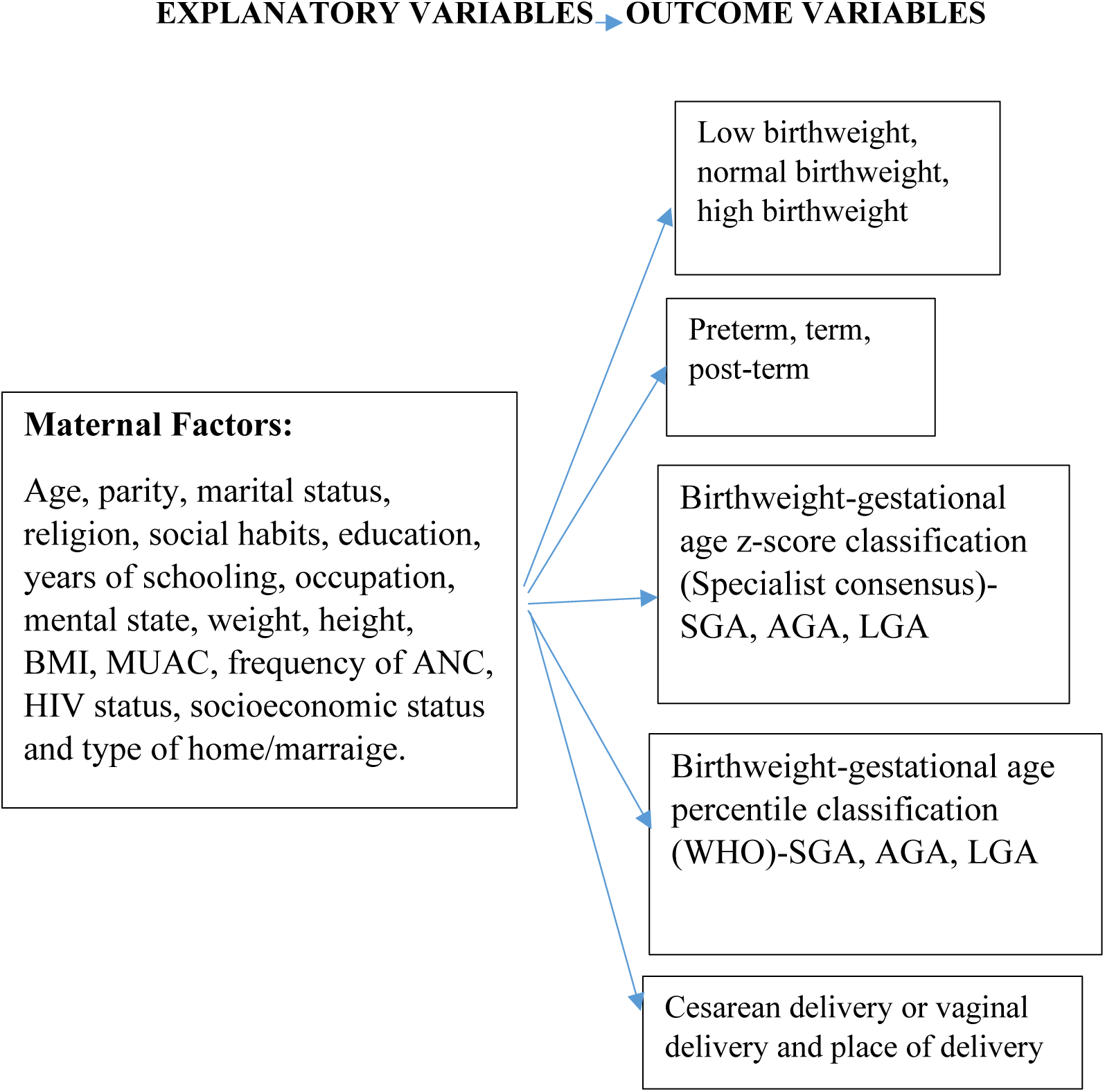
Concept of the study.

## Materials and methods

The methods employed in carrying out this study is discussed in this section.

### Study setting

Rwanda is a landlocked country located in central-eastern part of Africa. It is a low-income country with a population of about 13 million people. The country ‘health system is a pyramid with the top of pyramid, being the Ministry of health responsible for sector coordination and oversight, and setting health policies and strategies. Currently, Rwanda has 565 public health facilities including 504 Health centers, 7 Medicalised Health Centers, 40 District, 4 Provincial, 4 Specialized and 8 Referral hospitals. Health posts are entities working at lowest level and operating under public-private partnership models; there are 1222 Health Posts (HPs) in Rwanda.^13^

Non-availability of regular supplies of clean and safe water, has been a longstanding problem in Rwanda, as a whole, probably because of its landlocked and hilly terrains, making construction and supply of piped water, a major challenge. The public piped water flow infrequently and the taps and pipes may be rusted and breached in some places, especially in the rural areas, further leading to contamination of household water. Many families store rain water in big tanks for use in their homes and this may become polluted (in the writer’s opinion) because of difficulties of cleaning these storage tanks. A few non-profit organizations, such as USAID and Water-for-life have sunk bore holes in strategic locations in a few number of villages in the country, with the aim of alleviating this water problem.^13^

Gitwe village is located on a high altitude of 1,674 meters above sea level, in the southern province, 240km from Kigali, which is the capital city of Rwanda. Gitwe general hospital began in 1995, immediately after the genocide, for the purposes of providing medical services and later training to this isolated community. The hospital currently has 100% government support, since year 2020. The maximum number of deliveries at the hospital per month was about 200. Some of the challenges in the hospital include poor specialist coverage and few trained health workers, poor supply of equipment, water, electricity, laboratory services and medicines. Challenging cases are referred to the university of Rwanda teaching hospital in Butare or Kigali. Gitwe village was selected for this study, because there was no published birth data from this poorly researched, remote community. In 2019, birth, feeding and growth data on 529 healthy mother-singleton newborn pairs were compiled in this village, over a period of 12 months for this study, which was carried out in the delivery and postnatal wards of Gitwe general hospital and at its annex, the maternal and child health clinic.

### Data source and sample

This was a prospective cohort study design. Mother-newborn pairs were recruited consecutively, on first-come-first-serve basis. Maternal file review and newborn anthropometry [weight (kg), length (cm) and head circumference (cm) measurements, recorded to the nearest decimals] were carried out, soon after birth. Maternal sociodemographic, antenatal and nutritional data were obtained using questionnaires, which were read to the mothers and filled by the research assistants. Maternal anthropometry was measured at 6 weeks postpartum and recorded and their BMI and MUAC were computed. Four approaches were used to classify the newborn: gestational age, birthweight, birthweight-gestational age percentile (WHO), birthweight-gestational age z-score (Specialists consensus). Newborn gestational age was determined using the maternal last menstrual period (LMP), fetal ultrasound gestational age dating (preferably done at first trimester) and/or expanded new Ballard criteria and were classified into preterm, term and post-term. Birthweight regardless of gestational age, with cut-off points of 2.5-3.9kg for normal birth weight. Birthweight-gestational age newborn classification (globally recommended) into small-for-gestational age (SGA), appropriate-for-gestational age (AGA) and large-for-gestational age (LGA) was done: according to WHO using 10^th^-90^th^ percentiles cut-off points for AGA and according to Specialists consensus using ±2 z-score cut-off points for AGA. It is worth noting that newborn classification is rarely done routinely in the study site. Newborn place and mode of delivery were also recorded.

To ensure the quality of data collected, 2 registered nurses were trained as research assistants at Gitwe Hospital for 2 days on the over-all procedure of mother and newborn anthropometry and data collection by the investigator. The questionnaires were pre-tested before the actual data collection period, on 10 mother-infant pair participants (2% of the total sample). The investigator closely followed the day-to-day data collection process and ensured completeness and consistency of the questionnaires administered each day, before data entry.

### Statistical analysis

Data clean up, cross-checking and coding were done before analysis. These data were entered into Microsoft Excel statistical software for storage and then exported to SPSS version-26 for further analysis. Both descriptive and analytical statistical procedures were utilized. Participants’ categorical characteristics were summarised in frequencies and percentages. A multivariate, multinomial and binary logistic regression models and Chi test were created to examine the relationships between the antenatal maternal independent variables (age, parity, marital status, religion, social habits, education, years of schooling, occupation, mental state, weight, height, BMI, MUAC, frequency of ANC, HIV status, socioeconomic status and type of home/marraige) and dependent variables [gestational age, birthweight classification, birthweight-gestational age z-score classification (Specialists’ consensus), birthweight-gestational age percentile classification (WHO), place and mode of delivery] and to generate the odds ratio. Factors with p-values <0.1 were included in the regression models. Odds ratio (OR), with a 95% confidence interval (CI) were computed to assess the strength of association between independent and dependent variables. Significant environmental factors determining birth outcomes were cross-tabulated. For all, statistical significance was declared at p-value < 0.05. The reporting in this study were guided by the STROBE guidelines for observational studies [29].

### Ethics

Ethical approval from the Health Sciences Research Ethics Committee of the University of the Free State in South Africa (Ethical Clearance Number: UFS-HSD2018/1493/2901) was obtained. Written permission to collect data was obtained from the Director of Gitwe Hospital and the eligible mothers gave their informed consent before enrolment. The participants were given research identity numbers and the principal investigator was responsible for the safe keeping of the completed questionnaires and collected data, to ensure anonymity and confidentiality of the participants.

## Results

The following are the results obtained from the study.

### Participants

Five hundred and ninety-seven (597) babies were delivered at Gitwe Hospital, Rwanda, between 3^rd^ January and 9^th^ May 2019, out of which, eligible 529 mother-newborn pairs were enrolled into the study, Figure 2.

**Fig 2:**
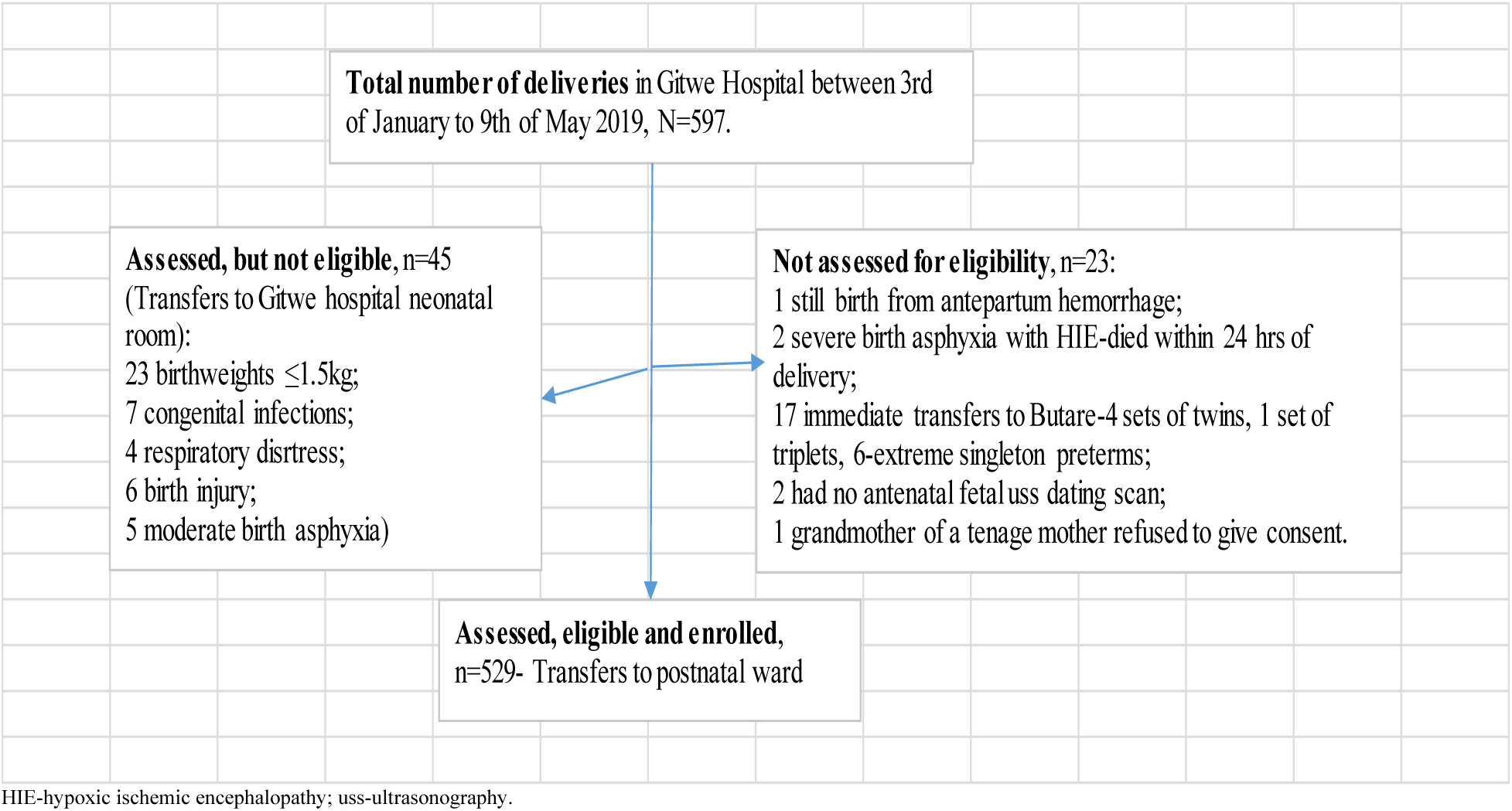
Flow of participants from admission to recruitment into study.

### Newborn characteristics

Prevalences of small for gestational age (SGA), appropriate for gestational age (AGA) and large for gestational age (LGA), were 21.4%, 71.6% and 7.0%, respectively, according to WHO percentile classification, Table 1. According to Specialists consensus z-score classification, there were SGA (5.3%), AGA (91.5%) and LGA (3.2%). Spearman rho correlation coefficient between the 2 classifications was fairly strong at 0.539. The majority were male (53.5%) and term babies (57.5%). Cesarean section delivery rate was 38.8% and 2 babies (0.4%) were not delivered in any health facility.

**Table 1:** Newborn characteristics (outcome variables) in the study.

| Characteristics | Frequency | Percent |
| --- | --- | --- |
| **Weight-for-gestational age (WHO, 1995)** |  |  |
| Small-for-gestational age (<10<sup>th</sup> percentile) | 113 | 21.4 |
| Appropriate-for-gestational age (10<sup>th</sup> -90<sup>th</sup> percentile) | 379 | 71.6 |
| Large-for-gestational age (>90<sup>th</sup> percentile) | 37 | 7.0 |
| **Birth weight (grams)** |  |  |
| Low birthweight (<2500) | 56 | 10.6 |
| Normal birthweight (2500-3999) | 461 | 87.1 |
| High birthweight (>3999) | 12 | 2.1 |
| **Mode of delivery** |  |  |
| Cesarean section | 205 | 38.8 |
| Vaginal delivery | 324 | 61.2 |
| **Place of delivery** |  |  |
| Outside health facility | 2 | 0.4 |
| Inside health facility | 527 | 99.6 |
| **Gender** |  |  |
| Female | 246 | 46.5 |
| Male | 283 | 53.5 |
| **Gestational age** |  |  |
| Preterm | 26 | 4.9 |
| Term | 304 | 57.5 |
| Post-term | 199 | 37.6 |
| **Total** | **529** | **100.0** |

### Maternal characteristics (independent variables)

The median age, parity, duration of schooling, weight, height and BMI of the mothers were 28 years, 2.0, 6 years, 62.0kg, 159cm and 24.7kg/m^2^, respectively, Table 2.

**Table 2:** Numerical characteristics of mothers in the study (n=529)

|  | Mean | SD | 95%CI | Median | IQR |
| --- | --- | --- | --- | --- | --- |
| Age of mother (years) | 27.9 | 6.44 | 27.33, 28.43 | 28.0 | 23, 33 |
| Parity of mother | 2.5 | 1.72 | 2.35, 2.64 | 2.0 | 1, 3 |
| Duration of schooling (years) | 7.4 | 3.33 | 7.10, 7.67 | 6.0 | 6, 9 |
| Weight (kg) | 63.2 | 8.72 | 62.50, 64.00 | 62.0 | 58, 68.5 |
| Height (cm) | 159.3 | 5.77 | 158.8, 159.8 | 159 | 157, 161 |
| BMI (kg/m<sup>2</sup>) | 24.9 | 3.09 | 24.6, 25.2 | 24.7 | 22.9, 26.6 |

The highest percentage of the mothers in the study, were young in age (43.9%), of primary school education (65.8%), unemployed/unskilled (58.8%), married (90.7%), christians (97.5%), middle class (75.0%), moderate parity (51.4%). Three (0.6%) mothers did not attend antenatal care clinic, and the majority (56.9%) had insufficient ANC visits (1-3). HIV positivity rate amongst the mothers was 5.1%. Majority of the mothers had good BMI (99.2%) and MUAC (95.3%), Table 3.

**Table 3:**
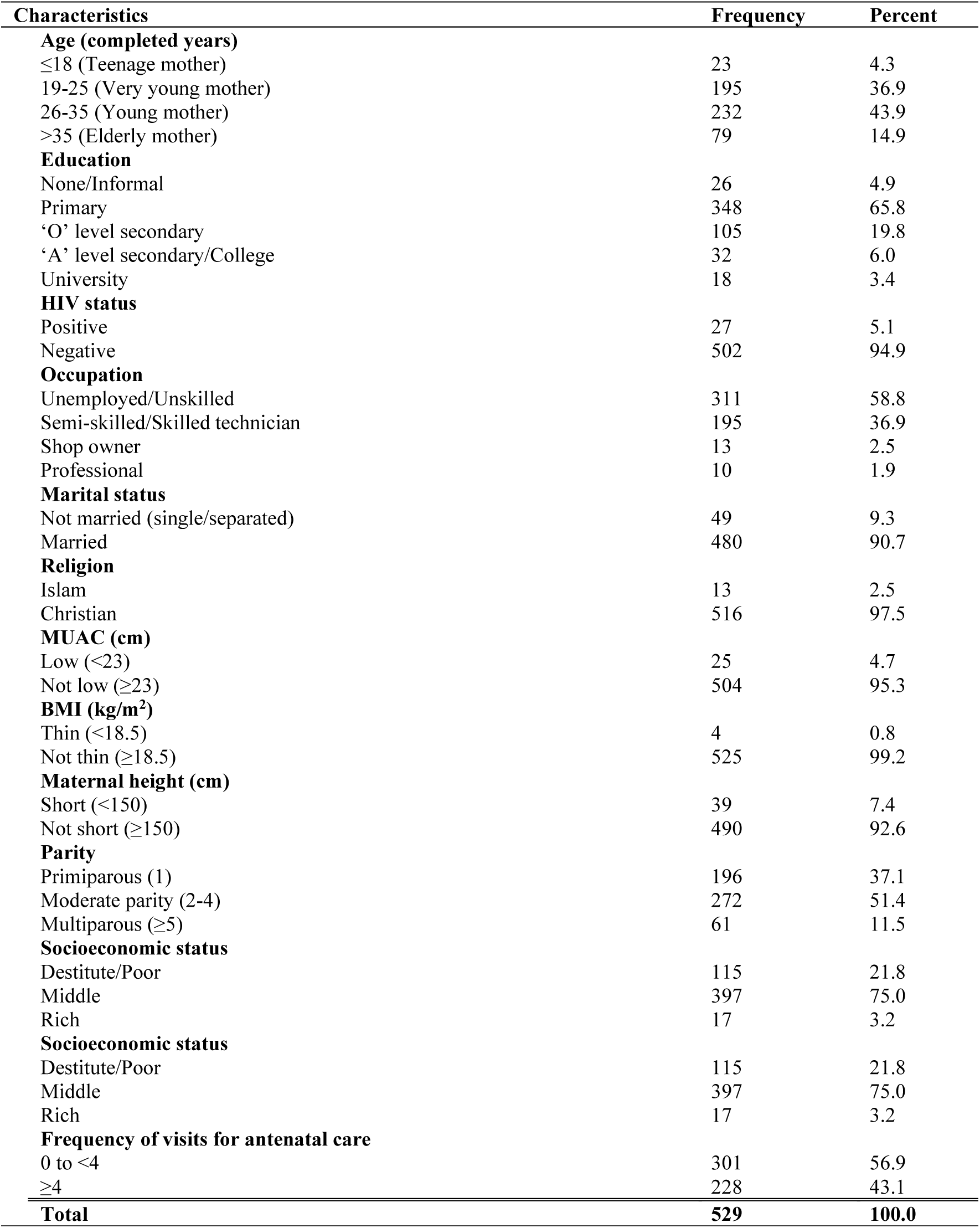
Categorical characteristics of mothers in the study (n=529)

| Characteristics | Frequency | Percent |
| --- | --- | --- |
| **Age (completed years)** |  |  |
| ≤18 (Teenage mother) | 23 | 4.3 |
| 19-25 (Very young mother) | 195 | 36.9 |
| 26-35 (Young mother) | 232 | 43.9 |
| >35 (Elderly mother) | 79 | 14.9 |
| **Education** |  |  |
| None/Informal | 26 | 4.9 |
| Primary | 348 | 65.8 |
| 'O' level secondary | 105 | 19.8 |
| 'A' level secondary/College | 32 | 6.0 |
| University | 18 | 3.4 |
| **HIV status** |  |  |
| Positive | 27 | 5.1 |
| Negative | 502 | 94.9 |
| **Occupation** |  |  |
| Unemployed/Unskilled | 311 | 58.8 |
| Semi-skilled/Skilled technician | 195 | 36.9 |
| Shop owner | 13 | 2.5 |
| Professional | 10 | 1.9 |
| **Marital status** |  |  |
| Not married (single/separated) | 49 | 9.3 |
| Married | 480 | 90.7 |
| **Religion** |  |  |
| Islam | 13 | 2.5 |
| Christian | 516 | 97.5 |
| **MUAC (cm)** |  |  |
| Low (<23) | 25 | 4.7 |
| Not low (≥23) | 504 | 95.3 |
| **BMI (kg/m<sup>2</sup>)** |  |  |
| Thin (<18.5) | 4 | 0.8 |
| Not thin (≥18.5) | 525 | 99.2 |
| **Maternal height (cm)** |  |  |
| Short (<150) | 39 | 7.4 |
| Not short (≥150) | 490 | 92.6 |
| **Parity** |  |  |
| Primiparous (1) | 196 | 37.1 |
| Moderate parity (2-4) | 272 | 51.4 |
| Multiparous (≥5) | 61 | 11.5 |
| **Socioeconomic status** |  |  |
| Destitute/Poor | 115 | 21.8 |
| Middle | 397 | 75.0 |
| Rich | 17 | 3.2 |
| **Socioeconomic status** |  |  |
| Destitute/Poor | 115 | 21.8 |
| Middle | 397 | 75.0 |
| Rich | 17 | 3.2 |
| **Frequency of visits for antenatal care** |  |  |
| 0 to <4 | 301 | 56.9 |
| ≥4 | 228 | 43.1 |
| **Total** | **529** | **100.0** |

### Multivariate logistic regression analysis of maternal factors predicting birth outcomes

According to the Wilks’ Lambda’s test in the multivariate logistic regression analysis in Table 4; significant maternal factors determining birth outcomes were: number of years spent in school (p=0.049), frequency of antenatal visits (p=0.021), educational level (p=0.011), HIV status (p=0.001), occupation (p=0.001) and marital status (p=0.007).

**Table 4:** Crosstabulation of significant maternal factors and mode of delivery, n=529.

|  | p-value | Mode of delivery |  | Total<br>(100.0) |
| --- | --- | --- | --- | --- |
|  |  | CS (%) | VD (%) |  |
| **Maternal BMI** | 0.008 |  |  |  |
| Thin (<18.5) |  | 1 (25.0) | 3 (75.0) | 4 |
| Normal (18.5-24.9) |  | 101 (34.1) | 195 (65.9) | 296 |
| Overweight (25-29.9) |  | 86 (42.4) | 117 (57.6) | 203 |
| Obese ($\geq 30$) | | 17 (65.4) | 9 (34.6) | 26 |
| **Frequency of antenatal care visits** | <0.001 |  |  |  |
| Low (<4) |  | 76 (25.2) | 225 (74.8) | 301 |
| Normal (4-6) |  | 121 (55.5) | 97 (44.5) | 218 |
| High (>6) |  | 8 (80.0) | 2 (20.0) | 10 |
| **Religion** | <0.001 |  |  |  |
| Moslem |  | 12 (92.3) | 1 (7.7) | 13 |
| Christian |  | 193 (37.4) | 32 (62.6) | 516 |
| **Education** | <0.001 |  |  |  |
| None/Informal education |  | 15(57.7) | 11 (42.3) | 26 |
| Primary |  | 108 (31.0) | 240 (69.0) | 348 |
| 'O' level secondary |  | 50 (47.6) | 55 (52.4) | 105 |
| 'A' level secondary/College |  | 20 (62.5) | 12 (37.5) | 32 |
| University |  | 12 (66.7) | 6 (33.3) | 18 |
| **Occupation** | <0.001 |  |  |  |
| Unemployed/Unskilled |  | 134 (43.1) | 177 (56.9) | 311 |
| Semi-skilled/Skilled technician |  | 55 (28.2) | 140 (71.8) | 195 |
| Entrepreneurs |  | 10 (76.9) | 3 (23.1) | 13 |
| Professional |  | 6 (60.0) | 4 (40.0) | 10 |
| **Maternal HIV status** | <0.001 |  |  |  |
| Positive |  | 24 (88.9) | 3 (11.1) | 27 |
| Negative |  | 181 (36.1) | 321 (63.9) | 502 |
| **Maternal Height** | <0.001 |  |  |  |
| <150cm (short) |  | 29 (74.4) | 10 (25.6) | 39 |
| $\geq 150$cm (not short) | | 176 (35.9) | 314 (64.1) | 490 |
| **Total** |  | **90 (17.0)** | **439 (83.0)** | **529(100.0)** |

### The tests of between-subjects multivariate regression post-hoc analysis

The tests of between-subjects effects analysis, revealed that the mode of delivery was significantly determined by maternal duration of schooling in years (p=0.001), weight (p=0.003), height (p=0.002), BMI (p=0.012), frequency of antenatal visits (p<0.001), educational level (p<0.001), occupation (p<0.001), HIV status (p<0.001) and religion (p=0.021).

The birthweight classification was significantly determined by maternal HIV status (p=0.027) occupation (p=0.031); marital status (p=0.024) and religion (p=0.047). Gestational age was significantly determined by marital status (p=0.002). Baby’s gender was significantly determined by maternal occupation (p=0.028). Newborn 1995 birthweight-gestational age percentile classification (WHO) was significantly determined by frequency of antenatal visits (p=0.010); while the classification using the 2007 birthweight-gestational age z-score was determined by parity (p=0.002).

### Binary logistic regression analysis investigating maternal factors predicting mode of delivery and neonatal gender

In the binary logistic regression analysis investigating the relationship between significant maternal factors (identified during multivariate logistic regression analysis) and mode of delivery, maternal level of education (wald 20.1), occupation (wald 18.7) and then frequency of antenatal visits (wald 17.2) were the strongest characteristics determining mode of delivery, among other significant factors such as number of years of schooling (wald 10.3), maternal weight (wald 5.4) and height (wald 7.0), BMI (wald 4.1), religion (wald 5.4) and HIV status (wald 7.4).

The lower the number of years of schooling, the less likely the mothers will deliver by cesarean section (p=0.001; OR=0.75; 95%CI 0.63, 0.90). Independent sample T-test result showed that the mean number of years of schooling in the group of women (205 in number, 38.8%), who delivered by c-section (8.13years, SD 4.12) was significantly higher than those (324 in number, 61.2%), who delivered vaginally (6.91years, SD 2.62), p<0.001 (95% CI of the difference was 0.64, 1.79). The primary school leavers were particularly, less likely to deliver by cesarean section (p=0.019; OR=0.02; 95%CI 0.001, 0.52) compared to their counterparts.

The professional group of mothers (p=0.023; OR=16.39; 95%CI 1.47, 100) were more likely to deliver by c-section compared to the unemployed or unskilled labour mothers. There was no longer any significant relationship (wald 1.9, p=0.163) between maternal occupation and sex of the newborn after binary logistic regression analysis. However, female babies were 1.84 (95% CI 0.78, 4.32) times more commonly observed with entrepreneur or professional mothers than their counterparts, who were either unemployed or engaged in an unskilled or technical kind of labour. Cross-tabulation between maternal occupation and prenatal exposure to potentially harmful social habits such as alcohol, herbs, tobacco revealed a significant relationship (p<0.001), in which, 66 (12.5%) out of the 529 mothers had prenatal exposure. Fifty-eight (87.9%) out of the 66 that were exposed were mothers that were either unemployed or engaged in unskilled labour. Eight (12.1%) of the 66 exposed, were mothers with semi-skilled or skilled technical jobs. Of note, was that, none of the entrepreneur or professional mothers were exposed. These unprescribed prenatal ingestions/exposures may have contained some sex selection drugs such as sex steroids.

The lower the maternal weight, the less likely the mother will deliver via cesarean section (p=0.020; OR 0.71; 95%CI 0.53, 0.95). Independent sample T-test result showed that the mean maternal weight in the group of women (205 in number, 38.8%), who delivered by c-section (64.19kg, SD 9.83) was significantly higher than those (324 in number, 61.2%), who delivered vaginally (62.59kg, SD 7.89), p=0.050 (95% CI of the difference was 0.00, 3.20).

The taller the mother, the more likely she will deliver via cesarean section (p=0.008; OR 1.38; 95%CI 1.09, 1.76). However, it was noted that the mean weight of the mothers, who were considered short (height <150cm) in the study, were significantly (p<0.001) lighter (57.74kg, SD 10.07) than those, that were considered tall (63.65kg, SD 8.46); 95%CI of the difference was - 8.71, -3.10. Independent sample T-test result showed that the mean maternal height in the group of women (205 in number, 38.8%), who delivered by c-section (158.76cm, SD 6.70) was similar to those (324 in number, 61.2%), who delivered vaginally (159.58cm, SD 5.07), p=0.135 (95% CI of the difference was -1.89, 0.26).

Independent sample T-test result showed that the mean number of antenatal visits in the group of women (205 in number, 38.8%), who delivered by c-section (3.86, SD 1.20) was significantly higher than those (324 in number, 61.2%), who delivered vaginally (3.19, SD 0.84), p<0.001 (95% CI of the difference was 0.48, 0.86).

The higher the maternal BMI, the more likely the mother will deliver via cesarean section (p=0.043; OR 2.11; 95%CI 1.02, 4.36). Independent sample T-test result showed that the mean maternal BMI in the group of women (205 in number, 38.8%), who delivered by c-section (25.43, SD 3.33) was significantly higher than those (324 in number, 61.2%), who delivered vaginally (24.58, SD 2.88), p=0.002 (95% CI of the difference was 0.32, 1.39).

Mothers of christian religion were less likely to deliver by c-section compared to the moslem mothers, p=0.020; OR=0.07 (95%CI 0.01, 0.66). HIV negative mothers were less likely to deliver by c-section, compared to their HIV positive counterparts, p=0.007, OR=0.17 (95%CI 0.05, 0.61).

### Multinomial logistic regression analysis of significant maternal factors determining neonatal birth size and gestational age

In the multinomial logistic regression analysis investigating the relationship between significant maternal factors (identified during multivariate logistic regression analysis) and neonatal birth weight, maternal marital status (wald 12.2, p<0.001) was the only characteristic that remained significant in determining neonatal birthweight, among other factors such as maternal HIV status (wald 1.5, p=0.226), religion (wald 0.3, p=0.577) and occupation (wald 1.5, p=0.221). The odds of a single mother giving birth to a LBW baby was 3.56 times higher (95%CI 1.75, 7.27) than that of a married mother. Likewise, the odds of a single mother giving birth to a preterm baby was 6.33 times higher (wald 10.2; p=0.001; 95%CI 2.04, 19.53) than that of a married mother.

The multiparous mother was less likely to give birth to an SGA (p=0.025, OR=0.17, 95%CI 0.04, 0.80) nor to an AGA (p=0.001, OR=0.17, 95%CI 0.06, 0.46) baby, compared to a mother with lower parity. The multiparous mother was more likely to give birth to a LGA baby, (according to birthweight-gestational age z-score classification). Number of antenatal visits was no longer significant in determining birthweight-for-gestational age percentile (according to WHO classification) after multinomial regression analysis (wald 1.1, p=0.288). However, those who had insufficient number of antenatal visits were more likely to deliver an SGA baby (OR=3.09; 95% CI 0.39, 28.82), compared to those, who received adequate antenatal care.

### Cross tabulation of significant maternal factors and birth outcomes

The obese mothers (17 out of 26, 65.4%), mothers who frequented the antenatal clinic more than 6 times during pregnancy (8 out of 10, 80.0%), moslem mothers (12 out of 13, 92.3%), those with university education (12 out of 18, 66.7%), entrepreneurs (10 out of 13, 76.9%), those who were HIV positive (24 out of 27, 88.9%) and those who were short (29 out of 39, 74.4%) were more likely to be delivered by c-section compared to their counterparts. Of note, the HIV positivity rate was significantly higher amongst the moslem mothers (3 out of 13, 23.1%) compared to the Christian mothers (24 out of 516, 4.7%), p=0.003. And a higher percentage of entrepreneurs were observed among the moslem (2 out of 13, 15.4%) compared to the christian mothers (11 out of 516, 2.1%)

In Table 5, a higher percentage of single mothers gave birth to LBW (13 out of 49, 26.5%) and preterm babies (6 out of 49, 12.2%) compared to their married counterparts. A higher percentage of primiparous mothers gave birth to SGA (14 out of 196, 7.1%) and the multiparous mothers gave birth to LGA babies (7 out of 61, 11.5%).

**Table 5:**
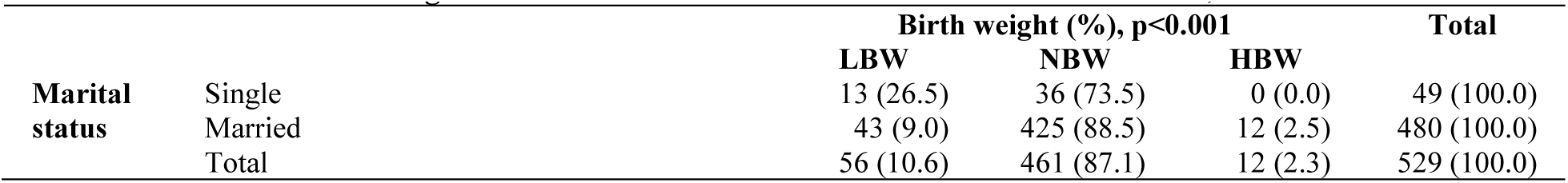
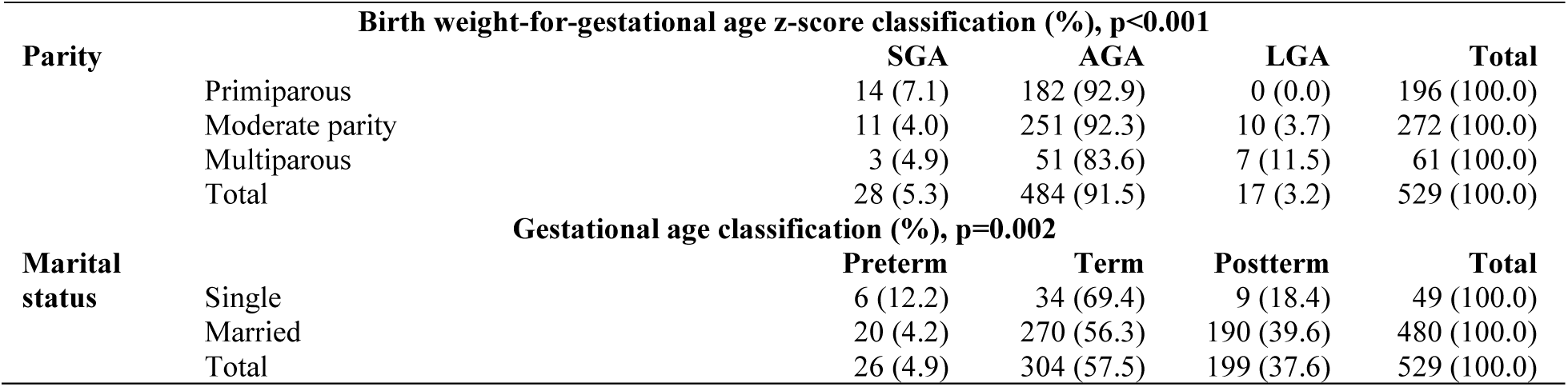
Crosstabulation of significant maternal factors and newborn characteristics, n=529.

## Discussion

Maternal predisposing factors to adverse neonatal outcomes are often times assumed to be similar in rural and urban settings. This assumption have led to many failed or failing interventions. According to WHO report of the 2019-2020 Rwanda DHS,^14^ the national rate of cesarean section was 15%; the rate was said to be higher in the urban areas (25%) and lower in the rural areas (13%); highest in Kigali city (25%) and lowest in the Northern province of Rwanda at 12%. Nineteen percent of the mothers with four or more ANC visits were delivered via cesarean section, as compared with only 3% of births to mothers with no ANC visits. Cesarean section deliveries were most common (19%) among first-order births and decrease as birth order increases. The proportion of cesarean section deliveries increases with increasing maternal education and wealth. In that same year of 2019, the rate of c-section was at least 38.8% (this rate could have been higher, if the very small and sick babies were included in this study) in this rural Gitwe village. This rate was even higher than the rates in Kigali city (RDHS 2020) and Namibia (16.1%), which was considered the highest in sSA according to Ahinkorah in 2022.^12^ This study finding is the reverse of what exists in literature, that state that c-section was more common in the urban than rural areas. The drivers of c-section were similar in both this study and in the RDHS (2020) report, except that maternal HIV positivity, excessive BMI and non-christian religion status were significant predictors of c-section delivery in this rural study. This was not the situation in the RDHS (2020) and Kibe et al (2022)’s reports.^9^ The World Health Organization (WHO) has recommended an optimal CS rate of 10–15%, above which, suggests overuse and of no benefit.^9^

Preterm and LBW births were significantly associated with unmarried women in this rural study. This was contrary to the findings of Rutayisire et al’s study,^15^ involving 817 women in 30 health facilities in 10 districts (including urban centres) of Rwanda; where married women (54.9%) were significantly associated with preterm births, compared with single mothers (7.1%). Preterm birth rate was 13.8% in Rutayisire et al’s study (2020), but was at least 4.9% in this Gitwe study. The wide disparity in the prevalence of preterm birth, may be as a result of excluding the very small and sick babies from this study and in the varying methods of determining gestational age in both studies. However, similar to Rutayisire et al’s study (2020), preterm birth was more common in the rural areas (74.3%) of Rwanda, compared to urban (25.7%).

LBW rate was 7% in the 2020 RDHS report, lower than the 10.6% reported in this rural (Gitwe) study. This study’s LBW rate was lower than the incidence of LBW (23.7%) reported in a tertiary hospital-based urban study in Ghana.^16^ Afaya et al. (2021) observed that being married has a protective effect on LBW [AOR = 0.60 (95%CI: 0.40–0.90), p = 0.013] compared to single mothers, similar to the findings in this study.^16^

The rate of SGA in a national retrospective study in Mexico (2021) was 6.7% and was noted to occur more among primiparous (aOR 1.42; 95% CI: 1.39, 1.43) mothers and mothers living in very high deprivation localities (aOR 1.39; 95% CI: 1.36, 1.43).^17^ This Mexico rate was at least 3 times lower than the 21.4% rate of SGA reported in this rural study. There was no information on SGA in the RDHS report.

Preterm, LBW and SGA births are the primary causes of perinatal illness and mortality worldwide.^18^ They are the biggest healthcare challenges as they are associated with long-term disability and financial strain from the costs of care especially, in underdeveloped nations. They have also long-term adverse effects, such as poor neurodevelopment leading to learning disabilities, cerebral palsy, and vision abnormalities among others.^19^

The exclusion of very small (<1.5kg), still births, major anomalies, very sick and multiple gestation babies, from the study (as resuscitation and transfer for specialized care were prioritized in these babies), could have weakened the study power and therefore was considered a limitation. Small numbers of preterm and post-term babies and some subcategories of mothers, such as the highly educated, professionals, entrepreneurs, mothers who did not use charcoal, mothers who did not attend ANC clinic; challenged our statistical analysis, thereby necessitating merging of some sub categories in some cases, during the analysis.

The strength of the study included the standard neonatal anthropometry and classification that were carried out for better management of these babies. This was not a routine practice in this non-specialised hospital. Birth data was generated for this remote community and made available for interested stakeholders to work with and improve the health status of mother and newborn in this community. This study also supplied additional information that was excluded in the RDHS.

## Conclusion and Recommendations

The magnitude of adverse neonatal outcomes in this rural study was unexpectedly higher than in the most recent Rwandan demographic health survey and those of several countries in the world. Therefore, mothers (not neglecting their marital, HIV, parity and religion status) residing in rural settings should be priortised on health intervention programs by healthcare providers, policymakers, and researchers, in order to lower the short-and long-term effects of these adverse birth outcomes.

## Data Availability

All data produced in the present study are available upon reasonable request to the authors

## Author Contributions

The corresponding author (Dr Ogah Adenike Oluwakemi) conceived and designed the study, collected data and conducted data analysis, interpreted the results, and drafted the manuscript. James Aaron Ogbole Oluwasegun reviewed and edited the manuscript for intellectual content. All the authors approved the final manuscript for submission.

## Acknowledgements

The authors are extremely grateful to the participants involved in this study, to the staff of Gitwe Hospital and clinic in Rwanda and to the research team. I am also indebted to my supervisors (Prof Andre Venter and Prof Corina Walsh), and my statistician Prof Gina Joubert, who analysed all the data for my PhD thesis.

